# Automated Diagnosis of COVID-19 Using Deep Learning and Data Augmentation on Chest CT

**DOI:** 10.1101/2020.04.24.20078998

**Authors:** Runwen Hu, Guanqi Ruan, Shijun Xiang, Minghui Huang, Qiaoyi Liang, Jingxuan Li

**Affiliations:** The School of Information Science and Technology, Jinan University, Guangzhou, 510632, China

**Keywords:** COVID-19, Chest CT, Deep Learning, Data Augmentation, Noisy Labels

## Abstract

**Background:** Coronavirus disease 2019 (COVID-19) has surprised the world since the beginning of 2020, and the rapid growth of COVID-19 is beyond the capability of doctors and hospitals that could deal in many areas. The chest computed tomography (CT) could be served as an effective tool in detection of COVID-19. It is valuable to develop automatic detection of COVID-19.

**Materials and Methods:** The collected dataset consisted of 1042 chest CT images (including 521 COVID-19, 397 healthy, 76 bacterial pneumonia and 48 SARS) obtained by exhaustively searching available data on the Internet. Then, these data are divided into three sets, referred to training set, validation set and testing set. Sixteen data augmentation operations are designed to enrich the training set in deep learning training phase. Multiple experiments were conducted to analyze the performance of the model in the detection of COVID-19 both in case of no noisy labels and noisy labels. The performance was assessed by the area under the receiver operating characteristic (AUC), sensitivity, specificity and accuracy.

**Results:** The data augmentation operations on the training set are effective for improvement of the model performance. The area under the receiver operating characteristic curve is 0.9689 with (95% CI: 0.9308, 1) in case of no noisy labels for the classification of COVID-19 from heathy subject, while the per-exam sensitivity, specificity and accuracy for detecting COVID-19 in the independent testing set are 90.52%, 91.58% and 91.21%, respectively. In the classification of COVID-19 from other hybrid cases, the average AUC of the proposed model is 0.9222 with (95%CI: 0.8418, 1) if there are no noisy labels. The model is also robust when part of the training samples is marked incorrectly. The average AUC is 92.23% in the case of noisy labels of 10% in the training set.

**Conclusion:** A deep learning model with insufficient samples can be developed by using data augmentation in assisting medical workers in making quick and correct diagnosis of COVID-19.

## Introduction

Coronavirus Disease 2019 (COVID-19), has surprised the world since the beginning of 2020 and is becoming a serious global concern. An infected patient may be leaded to acute respiratory distress or multiple organ failure. Due to its quick spread in the world, COVID-19 becomes a potentially life-threatening illness on the lives of billions of people [1–4]. To date (April sixteenth 2020), there have been two millions of confirmed cases all around the world. In many areas, the rapid growth of COVID-19 is beyond the capability of doctors and hospitals that could deal.

Although reverse-transcription polymerase chain reaction (RT-PCR) can confirm this disease with high specificity, the sensitivity of this diagnostic gold standard at the initial presentation of COVID-19 might not be high and suffer relatively long detection time [5, 6]. Meanwhile, ground glass infiltrates in chest computed tomography (CT) images, which have been described as representative of the COVID-19 [7,8], can be recognized by radiologists and experienced medical workers. Nine chest CT images of COVID-19 in mild, moderate and severe cases are shown in Fig. 1. In [9], the authors focus on interpreting the potential pathological basis from CT images and suggesting the future research and clinical directions, which will be greatly helpful for the radiologists in the clinical practice. Consequently, CT is an important tools in the diagnosis and treatment pathway of COVID-19 [10]. A CT scan takes only a few minutes, but some tens of minutes is needed for a group of experienced doctors to read the CT images and make the correct decision. In many areas, the virus spread quickly and there are a desperate shortage of experienced medical workers to evaluate large numbers of chest CT images. Therefore, how to break through the bottleneck of time of diagnosis and give the medical worker a hand in making the quick and correct decision becomes an import issue.

Artificial intelligence (AI) has made great success in many image application domains [11–13], such as in medical applications [14] by developing deep learning model in assisting doctors in reading CT images of lungs to detect cancer [15] and pneumonia in pediatric chest radiographs [16]. Toward this direction, AI-based automated image analysis on CT images is a potential solution for COVID-19. In the past two months, there were a few excellent works proposed for doctors in making efficient diagnosis of COVID-19 [17–19]. In [17], a convolutional neural network (CNN) using ResNet50 as the backbone to detect COVID-19 and distinguish it from community acquired pneumonia and other non-pneumonic lung diseases were proposed by using 4,356 3D chest CT exams. In [18], the authors developed an automated CT image analysis method to distinguish COVID-19 patients from those who do not have the disease by using deep learning on a testing set of 157 international patients. The two datasets in [17,18] are not open due to some reasons of privacy protection. Consequently, researchers tried to build public COVID-19 dataset in their researches [19, 20]. In [19], the authors built a public dataset consisting of 275 CT scans positive for COVID-19 and developed a CNN model using DenseNet169 on the dataset. The dataset in [20] is composed of a number of COVID-19 chest X-rays or CT scans collected for computer analysis.

Usually, the performance of AI methods, especially based on deep learning, depend on a large and reliable dataset. However, a large and reliable dataset is considered to be one of the biggest challenges for developing deep learning method in medical applications [14], especially for current AI-based diagnosis of COVID-19. Firstly, COVID-19 samples are difficult to be collected because of protection of patient privacy. Besides, building a reliable dataset from clinical data needs experts to annotate labels. Since the complexity of the medical data and the different experiences among the experts, a medical dataset whose labels are completely reliable is not available, meaning that there may have noisy (or incorrect) labels in some training samples. In this situation, data augmentation (DA) technique could be used to generalize an AI model based on deep learning and the model has resistance to noisy labels in some training samples is required.

We proposed a new COVID-19 diagnosis method by using CNN with ShuffleNet V2 as the backbone so as to efficiently distinguish the COVID-19 patients from those who are not infected or infected by other pneumonia (bacterial pneumonia or SARS). We built a new dataset (which can be downloaded from the website: https://github.com/KevinHuRunWen/COVID-19) for the testing. The purposes are to investigate whether AI model based on deep learning with DA technique can be developed while initially the number of COVID-19 samples is not enough to train deep learning algorithms, and the robustness of the AI model in the case of noisy labels in the training samples. This is useful for doctors in analyzing potentially large numbers of chest CT exams on diagnosis of COVID-19 as early as possible.

## Materials and Methods

### Materials

Considering about ethical issues such as the privacy of patients, the accessible chest CT images of healthy persons and patients with COVID-19 are seriously inadequate, and therefore, we strove and spent a lot of energy to collect the CT images from several different open sources and innovative papers. Most of the CT images are obtained from the work [19] and the online site at https://www.sirm.org/category/senza-categoria/covid-19/ while the rest samples are from [20]. Since the wide range of data sources may cause the problem of repetition or high correlation among the chest CT images, small part of collected CT images in these data sources are discarded. As a result, the chest CT images in the new dataset consist of 521 COVID-19 exams, 397 healthy subjects, 76 bacterial pneumonia and 48 SARS cases. In the total 521 COVID-19 exams, 349 are downloaded from [19], 13 images are obtained from [20] and 159 are from the online site above. Based on these collected samples, the training set, validation set and testing set are allocated as shown in Table 1 by considering the samples in number. In Table 1, there are 313 chest CT images with COVID-19 and 313 hybrid chest CT images (242 healthy chest CT images, 46 bacterial pneumonia images and 28 SARS images) in the training set, 104 chest CT images with COVID-19 and 104 hybrid chest CT images (83 healthy chest CT images, 15 bacterial pneumonia images and 10 SARS images) in the validation set, and 104 chest CT images with COVID-19 and 104 hybrid chest CT images (72 healthy chest CT images, 15 bacterial pneumonia images and 10 SARS images) in the testing set.

**Fig. 1.**
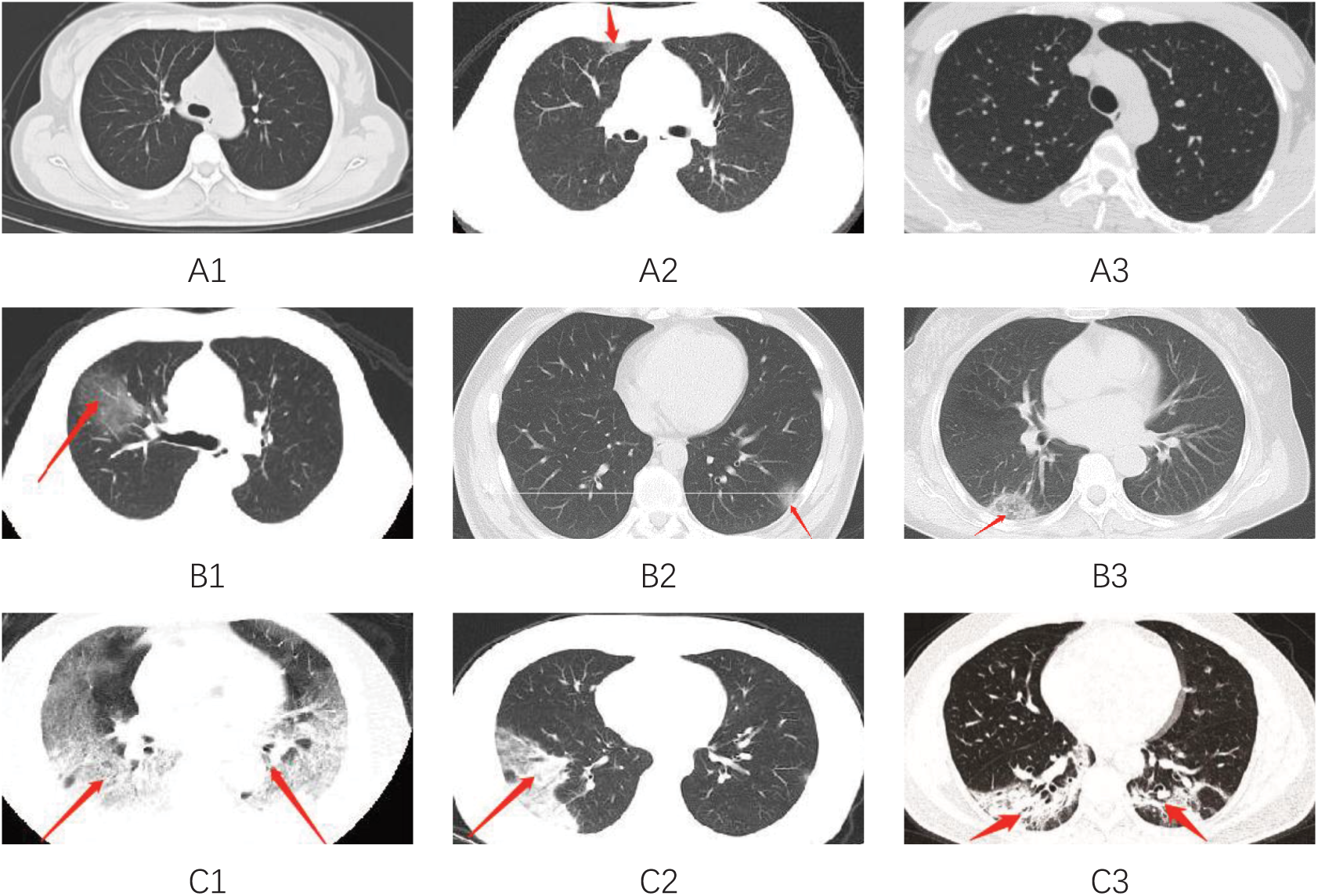
Demonstrations on chest CT images of COVID-19. A1-A3, mild pneumonia patient showed patchy ground-glass opacity with clear borderline in a transverse chest CT image; B1-B3, moderate patients with large ground-glass opacity and some with ambiguous borderline; and C1-C3, severe cases were characterized by typical white lung change as high-density mass shadows and multiple lobular consolidations were observed.

**Table 1.** Numbers of the samples in training, validation and testing sets.

| Types | Training set | Validation set | Testing set | Total |
| --- | --- | --- | --- | --- |
| COVID-19 | 313 | 104 | 104 | 521 |
| Healthy | 242 | 83 | 72 | 397 |
| Bacterial | 46 | 15 | 15 | 76 |
| SARS | 28 | 10 | 10 | 48 |

Until now, it is still difficult to collect enough and high-quality COVID-19 chest CT images for the AI model. Usually, the AI-based methods especially for deep learning, need enough data for training and testing, otherwise it will be easily prone to the overfitting problem, and inhibit its capability to generalize to unseen invariant data [21]. A solution is to use DA operations [22–25] to enrich the training samples for generalization and robustness of the model when the dataset is till small. In our method, the training set is expanded by using five types of DA techniques for improvement of the AI model while keeping the other two sets unchanged.

### AI Model Based on ShuffleNet V2

Considering the accuracy and running speed, we constructed our AI model based on the ShuffleNet V2 network [26, 27]. The CNN model using ShuffleNet V2 as the backbone is the most advanced lightweight model available, which has better accuracy and run faster than previous lightweight networks with the same computation condition. The input of ShuffleNet V2 network is a chest CT image in a time after performing a random cropping operation. The output of the ShuffleNet V2 network is then fed to a linear layer to generate the diagnostic result. With enough data, the network can learn to recognize the characteristics of COVID-19, and extract the feature which will be sent to the linear layer to get a probability score for diagnosis of COVID-19. The structure of our model is shown in Fig. 2. When the AI model is trained, the diagnosis result will be immediately obtained just like experienced clinician but faster.

**Fig. 2.**
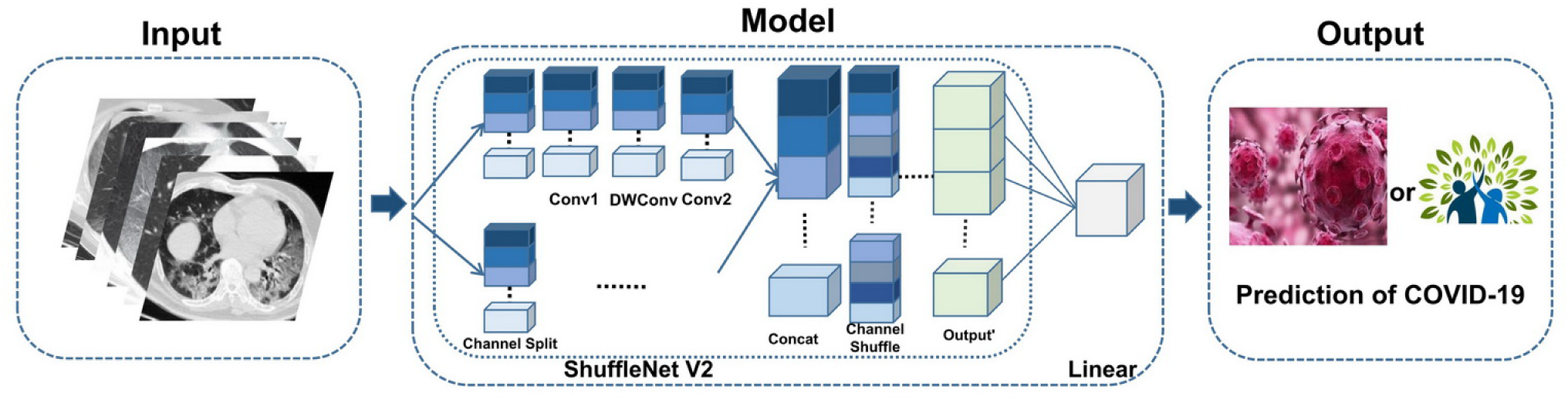
Flow diagram of the proposed AI method.

### Statistical Analysis

Containing the true value under a certain probability, confidence interval (CI) is the estimation interval of true parameters constructed by sample statistics. We adopt 95% confidence interval (95%CI) as a significant methd to evaluate the performance of our model on the validation and testing sets. With the help of 95%CI, the confidence interval calculated from the datasets we used is 95% likely to contain the real value. The expression of 95% confidence interval is simplified to “[95%CI: *A*, *B*]” in following sections, where *A* and *B* represent the lower bound and the upper bound of the interval, respectively.

## Data Augmentation

The section discuss five types of DA techniques including sixteen operations used for the training set at first. This is followed by an experimental evaluation on the effect of the DA operations on model performance.

### Augmentation Operations

Empirically, over one thousand images are needed in a class in image classification. As listed in Table 1, the dataset is till not enough for development of the proposed model. As shown in Table 2, we adopt five types of DA techniques including 16 manipulations in total on the samples in the training set. For each sample in the training set, we can get 16 new variants marked with the same label as the original one. In Fig. 3, the visual effect of five typical DA operations on the original sample have been plotted.

**Table 2.** Types of Data Augmentation.

| Data Augmentation Types | Parameters | Count |
| --- | --- | --- |
| Flip | Flip (vertical/horizontal/vertical +horizontal) | 3 |
| Rotation | Rotate(-90,60,90,150) | 4 |
| Translation | Translation(10,50) | 1 |
| Brightness Adjustment | Brightness(0.5/1.5) | 2 |
| Flip+Brightness Adjustment | Flip(vertical/horizontal/vertical +horizontal)+Brightness(0.5/1.5) | 6 |

**Fig. 3.**
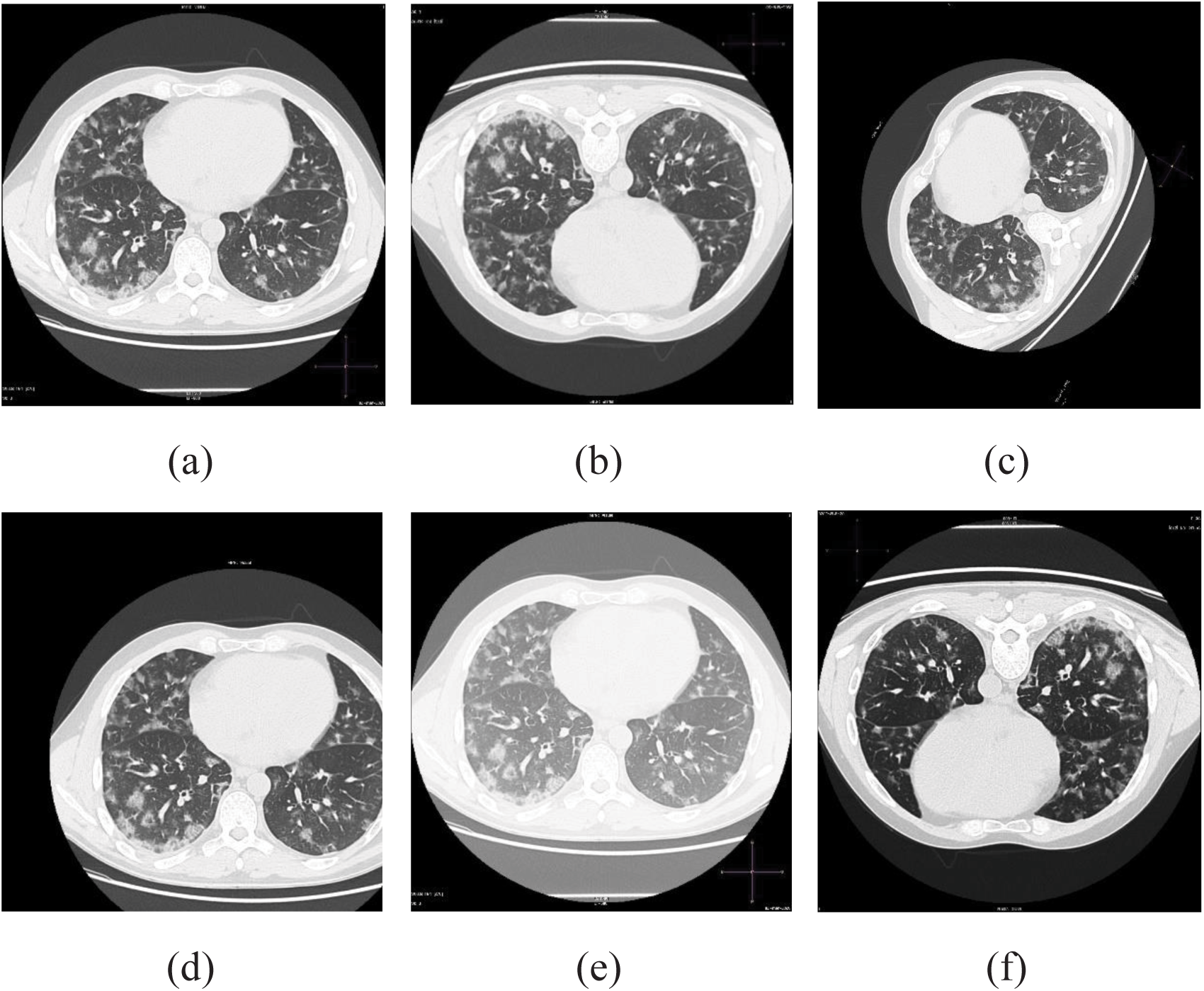
CT Images of lungs of a COVID-19 case and its five DA versions. (a) the original CT image, (b) flip vertically, (c) rotation of anticlockwise 60 degrees, (d) translation of 10 rows and 50 columns, (e) 1.5 times brightness, (f) 0.5 times brightness and flip vertically and horizontally.

### Experimental Evaluation

The samples in the training set, validation set and testing set have been allocated as shown in Table 1. Totally 555 samples in the training set (including 313 COVID-19 exams and 242 healthy healthy subjects) are used to train the model by the validation set (104 COVID-19 and 83 healthy) and then test the model performance by the testing set (104 COVID-19 and 72 healthy). In order to verify the validity of the DA operations, we conducted two experiments to evaluate the effectiveness of the DA operations by using the original training set and the expanded training set with the 16 DA operations listed in Table 2, respectively.

In the first experiment the model has been trained by the original training set, the loss values by the validation set are plotted in Fig. 4 with *blue* line. The other experiment is to use the expanded training set while keeping the validation set and the testing set unchanged. After performing all the 16 DA operations on the the samples (313 COVID-19 and 242 healthy) in the training set, we obtained 9,435 chest CT images in the expanded training set in total while keeping the samples in the validation and testing sets unchanged. The loss values of the experiment are plotted in Fig. 4 with *red* line. We can see from the figures in Fig. 4 that the performance of the model has been improved to some extent by using DA techniques on the training set. After the DA, the loss function is more stable and the values are smaller than the original training set.

**Fig. 4.**
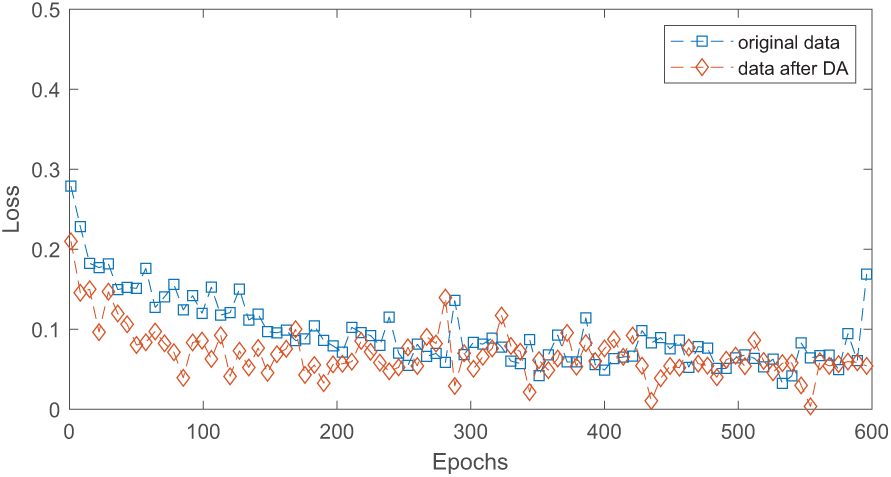
The loss functions of the two developed models with the original training set and the expanded one by using DA.

Also, we can see from Fig. 4 the AI model becomes stable after running 200 epoches and keeps well stable between 400 and 600 epoches. Consequently, we chose the models in the epoch period between 400 and 600 to compute the results of the model in the testing set in statistical way, as listed in Table 3. From this table, we can see that the model developed with the expanded training set has higher sensitivity, specificity and AUC in average while it is with a smaller average loss value. Take AUC metric as an example. Before the DA operations, the mean value and the variance are 0.8847 and 0.006, respectively. After the DA, the mean is up to 0.9222 while the variance is down to 0.0017. As for the loss values, the model with the expanded training set output a smaller mean and a smaller variance, from 0.1620 and 0.1122 down to 0.0155 and 0.0013, respectively. These results show the DA operations used in the paper has an ability in improving the model performance by expanding the samples in the training set when the samples are not enough.

**Table 3.** Averages and variances of the loss, sensitivity, specificity and AUC values in the range [400, 600] on the the testing set.

| Value | Mean |  | Variance |  |
| --- | --- | --- | --- | --- |
|  | Original | DA | Original | DA |
| Loss | 0.1620 | 0.1122 | 0.0155 | 0.0013 |
| Sensitivity | 0.8251 | 0.8571 | 0.030 | 0.0075 |
| Specificity | 0.7790 | 0.8488 | 0.0311 | 0.0099 |
| AUC | 0.8847 | 0.9222 | 0.0062 | 0.0017 |

## Results

We conduct a number of retrospective experiments to report the model performance. In the following experiments, we choose those epochs ranged from 400 to 600 as a stable period computed in the training process by the training and validation sets. Instead of randomly using a model in the stable period, we use the average of the models in the stable period on the testing set to report the model performance on the testing set. The basic reason is that the best performance in the validation set is usually not corresponding to the best rest results in the testing set. With the consideration of noisy labels, we distinguish COVID-19 from Healthy Cases at first. This is followed by a classification of COVID-19 from other cases. Experimental results show that the proposed AI model has an ability for diagnosis of COVID-19.

### Classification of COVID-19 from Healthy Cases

We have 521 COVID-19 and 397 healthy subjects in the classification testing. The samples are allocated to training, validation and testing sets by referring to Table 1. After performing all the 16 DA operations on the the samples (313 COVID-19 and 242 healthy) in the training set, we obtained 9,435 chest CT images in the expanded training set in total while keeping the samples in the validation set (104 COVID-19 and 83 healthy) and the testing set (104 COVID-19 and 72 healthy) unchanged. In the case of no noisy labels, experimental results obtained by the validation set are plotted in Fig. 5. We can see from the figures in Fig. 5 that the model is going to be stable in the all four performance metrics (loss value, sensitivity, specificity and AUC). We adopt the epochs between 400 and 600 as the stable period, and list the performance of the proposed model in classifying COVID-19 from healthy lungs under different noise fractions (NFs) in both the validation and testing sets, as shown in Table 4. In the table marked with *bold* font, the average sensitivity and specificity in the stable period are 90.52% and 91.58%, respectively. The corresponding average AUC value is 0.9689 with [95%CI: 0.9308, 1] while the average accuracy is 91.21%.

**Fig. 5.**
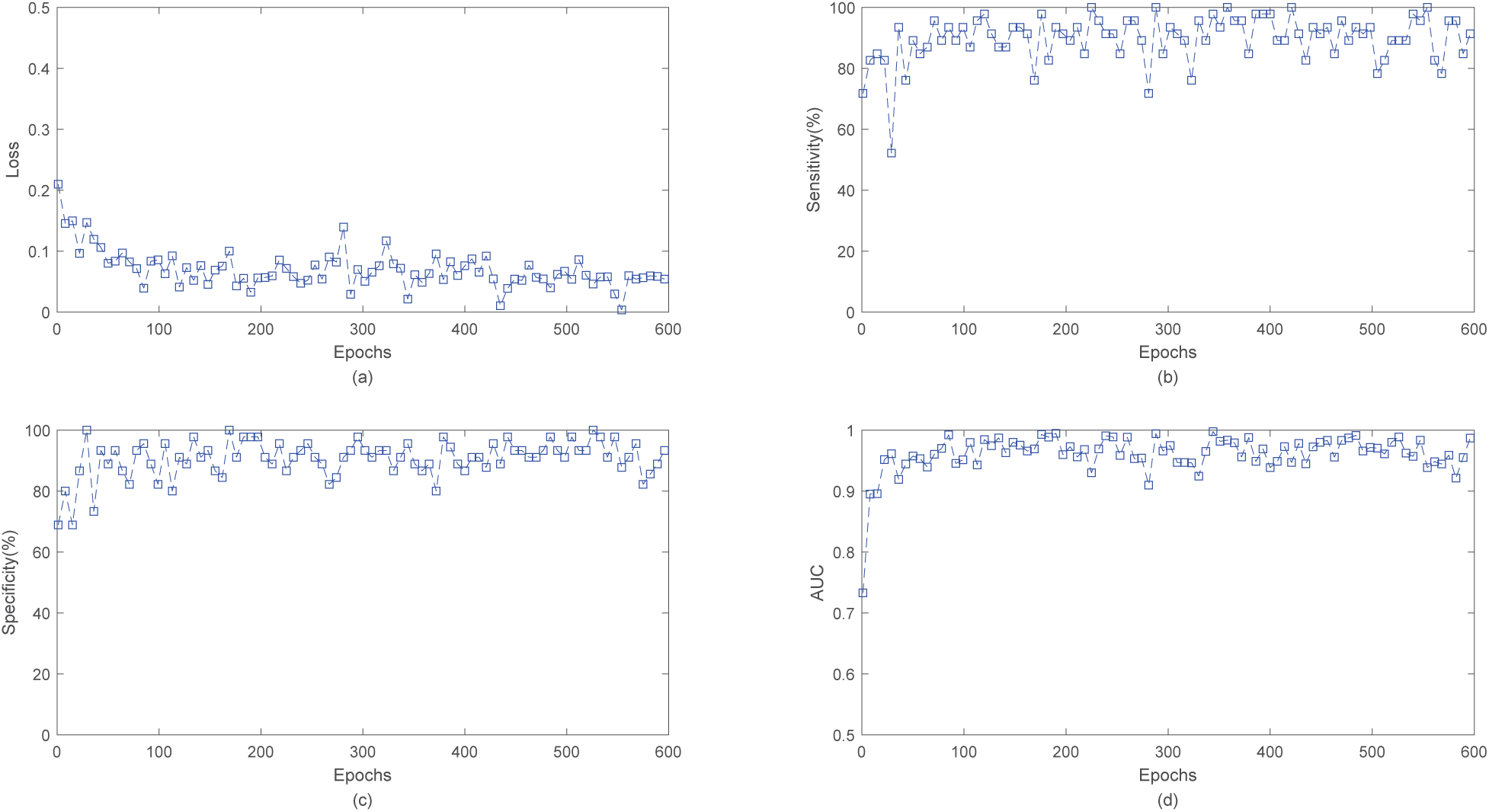
Model performance in case of no noisy labels. The loss, sensitivity, specificity and AUC are plotted in subfigures (a), (b), (c) and (d), respectively.

**Table 4.** Model performance in classifying the COVID-19 and healthy lungs under different noise fractions (NFs) in both the validation and testing sets.

| Arguments |  | Validation set |  |  | Testing set |  |  |
| --- | --- | --- | --- | --- | --- | --- | --- |
| Types | NFs(%) | Max | Min | Mean | Max | Min | Mean |
| Accuracy(%) | 0 | 92.59 | 70.37 | 86.97 | 97.92 | 76.64 | **91.21** |
|  | 1 | 91.67 | 62.96 | 85.43 | 97.92 | 76.34 | 89.06 |
|  | 5 | 90.74 | 73.15 | 82.52 | 94.79 | 76.34 | 85.82 |
|  | 10 | 87.04 | 61.11 | 78.60 | 93.75 | 73.51 | 83.91 |
|  | 15 | 82.41 | 60.19 | 73.62 | 84.67 | 68.90 | 76.85 |
|  | 20 | 76.85 | 54.63 | 66.25 | 82.29 | 61.61 | 72.88 |
|  | 25 | 75.00 | 51.85 | 65.19 | 81.25 | 56.40 | 69.66 |
|  | 30 | 77.78 | 53.70 | 65.76 | 79.17 | 56.99 | 68.39 |
|  | 40 | 68.52 | 40.74 | 56.95 | 69.64 | 44.94 | 57.90 |
| Sensitivity(%) | 0 | 98.00 | 70.00 | 89.80 | 100.00 | 54.35 | **90.52** |
|  | 1 | 100.00 | 68.00 | 88.99 | 100.00 | 67.39 | 91.31 |
|  | 5 | 98.00 | 60.00 | 86.06 | 97.83 | 60.87 | 83.00 |
|  | 10 | 100.00 | 60.00 | 82.96 | 100.00 | 58.70 | 85.52 |
|  | 15 | 98.00 | 46.00 | 75.32 | 91.30 | 41.30 | 75.50 |
|  | 20 | 94.00 | 42.00 | 68.30 | 97.83 | 36.96 | 69.66 |
|  | 25 | 86.00 | 42.00 | 65.70 | 89.13 | 39.13 | 66.95 |
|  | 30 | 90.00 | 44.00 | 67.17 | 97.83 | 45.65 | 68.81 |
|  | 40 | 88.00 | 34.00 | 62.07 | 91.30 | 43.48 | 65.42 |
| Specificity(%) | 0 | 100.00 | 46.94 | 83.73 | 100.00 | 30.00 | **91.58** |
|  | 1 | 97.96 | 36.73 | 81.79 | 100.00 | 53.33 | 85.55 |
|  | 5 | 100.00 | 61.22 | 79.04 | 100.00 | 64.44 | 88.63 |
|  | 10 | 89.80 | 36.73 | 74.40 | 100.00 | 51.11 | 82.31 |
|  | 15 | 85.71 | 46.94 | 71.82 | 97.78 | 48.89 | 78.53 |
|  | 20 | 95.92 | 34.69 | 65.17 | 97.78 | 37.78 | 76.23 |
|  | 25 | 87.76 | 38.78 | 64.95 | 97.78 | 44.44 | 72.36 |
|  | 30 | 85.71 | 40.82 | 64.22 | 91.11 | 37.78 | 67.93 |
|  | 40 | 75.51 | 20.41 | 51.49 | 84.44 | 22.22 | 50.22 |
| AUC | 0 | 0.9796 | 0.8514 | 0.9293 | 0.9995 | 0.8812 | **0.9689** |
|  | 1 | 0.9722 | 0.6461 | 0.9251 | 0.9971 | 0.8150 | 0.9608 |
|  | 5 | 0.9649 | 0.8257 | 0.9002 | 0.9845 | 0.8643 | 0.9396 |
|  | 10 | 0.9392 | 0.7392 | 0.8611 | 0.9860 | 0.7947 | 0.9223 |
|  | 15 | 0.8767 | 0.6816 | 0.8033 | 0.9377 | 0.7643 | 0.8486 |
|  | 20 | 0.7988 | 0.6273 | 0.7075 | 0.8899 | 0.7005 | 0.7993 |
|  | 25 | 0.7804 | 0.6033 | 0.6929 | 0.8589 | 0.6314 | 0.7628 |
|  | 30 | 0.8314 | 0.5694 | 0.7092 | 0.8435 | 0.5961 | 0.7436 |
|  | 40 | 0.6918 | 0.4400 | 0.5846 | 0.7449 | 0.4556 | 0.5823 |

### Classification of COVID-19 from Other Cases

In practical diagnosis of COVID-19, there may have the other kind of pneumonia, such as bacterial pneumonia and SARS, which needed to be distinguished from COVID-19. In order to test the ability of the model to classify COVID-19 from other cases, in the experiment we use the dataset including 521 COVID-19 images and 521 hybrid chest CT images consisting of 397 healthy, 76 bacterial and 48 SARS cases. The samples are allocated to training, validation and testing sets by referring to Table 1. Similarly, we expand the training set and test the model performance, as listed in Table 5. In this table marked with *bold* font, the average sensitivity and specificity in the stable period are 85.71% and 84.88%, respectively. The corresponding average AUC value is 0.9222 with [95%CI: 0.8418, 1] while the average accuracy is 85.40%.

**Table 5.** Performance of the proposed model in classifying the COVID-19 and the other pneumonia under different noise fractions (NFs) in both the validation and testing sets.

| Arguments |  | Validation set |  |  | Testing set |  |  |
| --- | --- | --- | --- | --- | --- | --- | --- |
| Types | NF(%) | Max | Min | Mean | Max | Min | Mean |
| Accuracy(%) | 0 | 90.74 | 59.72 | 81.52 | 93.06 | 69.44 | **85.40** |
|  | 1 | 90.74 | 61.11 | 81.18 | 91.67 | 62.04 | 84.89 |
|  | 5 | 87.04 | 63.89 | 78.50 | 89.81 | 65.74 | 82.61 |
|  | 10 | 82.87 | 62.96 | 75.44 | 87.04 | 66.67 | 79.11 |
|  | 15 | 79.63 | 61.57 | 72.79 | 85.19 | 64.35 | 76.32 |
|  | 20 | 79.63 | 62.04 | 71.15 | 80.09 | 64.35 | 72.46 |
|  | 25 | 75.93 | 60.19 | 67.33 | 76.85 | 59.72 | 68.64 |
|  | 30 | 69.44 | 51.85 | 59.86 | 75.00 | 56.02 | 67.48 |
|  | 40 | 68.52 | 46.30 | 57.50 | 66.20 | 50.00 | 58.19 |
| Sensitivity(%) | 0 | 98.08 | 48.08 | 82.94 | 99.04 | 42.31 | **85.71** |
|  | 1 | 95.19 | 35.58 | 81.66 | 98.08 | 29.81 | 85.13 |
|  | 5 | 95.19 | 32.69 | 80.74 | 100.00 | 34.62 | 84.29 |
|  | 10 | 96.15 | 28.85 | 79.79 | 98.08 | 35.58 | 83.87 |
|  | 15 | 93.27 | 31.73 | 76.08 | 97.12 | 29.81 | 79.93 |
|  | 20 | 95.19 | 50.00 | 80.36 | 95.19 | 41.35 | 78.72 |
|  | 25 | 88.46 | 40.38 | 67.70 | 90.38 | 38.46 | 69.67 |
|  | 30 | 84.62 | 28.85 | 62.58 | 94.23 | 40.38 | 71.60 |
|  | 40 | 89.42 | 23.08 | 56.87 | 87.50 | 25.00 | 60.80 |
| Specificity(%) | 0 | 94.23 | 22.12 | 79.93 | 99.04 | 25.96 | **84.88** |
|  | 1 | 97.12 | 30.77 | 80.63 | 98.08 | 38.46 | 84.73 |
|  | 5 | 96.15 | 38.46 | 76.30 | 98.08 | 37.50 | 80.88 |
|  | 10 | 98.08 | 40.38 | 71.24 | 95.19 | 43.27 | 74.38 |
|  | 15 | 95.19 | 36.54 | 69.48 | 96.15 | 36.54 | 72.74 |
|  | 20 | 94.23 | 30.77 | 61.85 | 94.23 | 35.58 | 66.21 |
|  | 25 | 91.35 | 38.46 | 66.75 | 91.35 | 37.50 | 67.66 |
|  | 30 | 81.73 | 25.00 | 57.07 | 85.58 | 31.73 | 63.33 |
|  | 40 | 84.62 | 29.81 | 58.14 | 83.65 | 27.88 | 55.64 |
| AUC | 0 | 0.9490 | 0.6661 | 0.8903 | 0.9778 | 0.6766 | **0.9222** |
|  | 1 | 0.9435 | 0.7346 | 0.8896 | 0.9696 | 0.6804 | 0.9162 |
|  | 5 | 0.9286 | 0.7243 | 0.8732 | 0.9705 | 0.7794 | 0.9024 |
|  | 10 | 0.8939 | 0.7519 | 0.8349 | 0.9374 | 0.7905 | 0.8652 |
|  | 15 | 0.8723 | 0.7060 | 0.7995 | 0.9038 | 0.7497 | 0.8398 |
|  | 20 | 0.8601 | 0.7036 | 0.7808 | 0.8645 | 0.7077 | 0.7966 |
|  | 25 | 0.7936 | 0.6565 | 0.7302 | 0.8215 | 0.6515 | 0.7424 |
|  | 30 | 0.7605 | 0.4897 | 0.6127 | 0.8043 | 0.5980 | 0.7192 |
|  | 40 | 0.6917 | 0.4701 | 0.5836 | 0.6870 | 0.4996 | 0.5882 |

### Effect of Noisy Labels

The complexity of medical resource and the differences among medical worker in clinical experiences have an important effect on whether a chest CT image is labeled correctly. In other words, small part of samples could be marked incorrectly in the dataset. Toward the noisy label problem, we test and observe the robustness of the model to noise fraction. With a noise fraction, the corresponding samples in proportion will be randomly selected from the training sets and marked with incorrect labels. These noisy samples will keep unchanged in the whole training process. When the model runs stable by referring to the validation set, we can get the corresponding stable period. Fig.6 plots the loss values in the validation set under different noise fractions. In Fig.6(a), the loss function becomes stable in the range between 200 to 600 epochs. From Fig.6(b) to Fig.6(i), we can observe that when there exist noisy labels, the loss functions of the validation set need more epoches to reach a stable state. The larger the noise fraction is, the more epochs are required for the model training. This explains why we choose the epoches ranged from 400 to 600 as the stable period to report the results on the testing set.

**Fig. 6.**
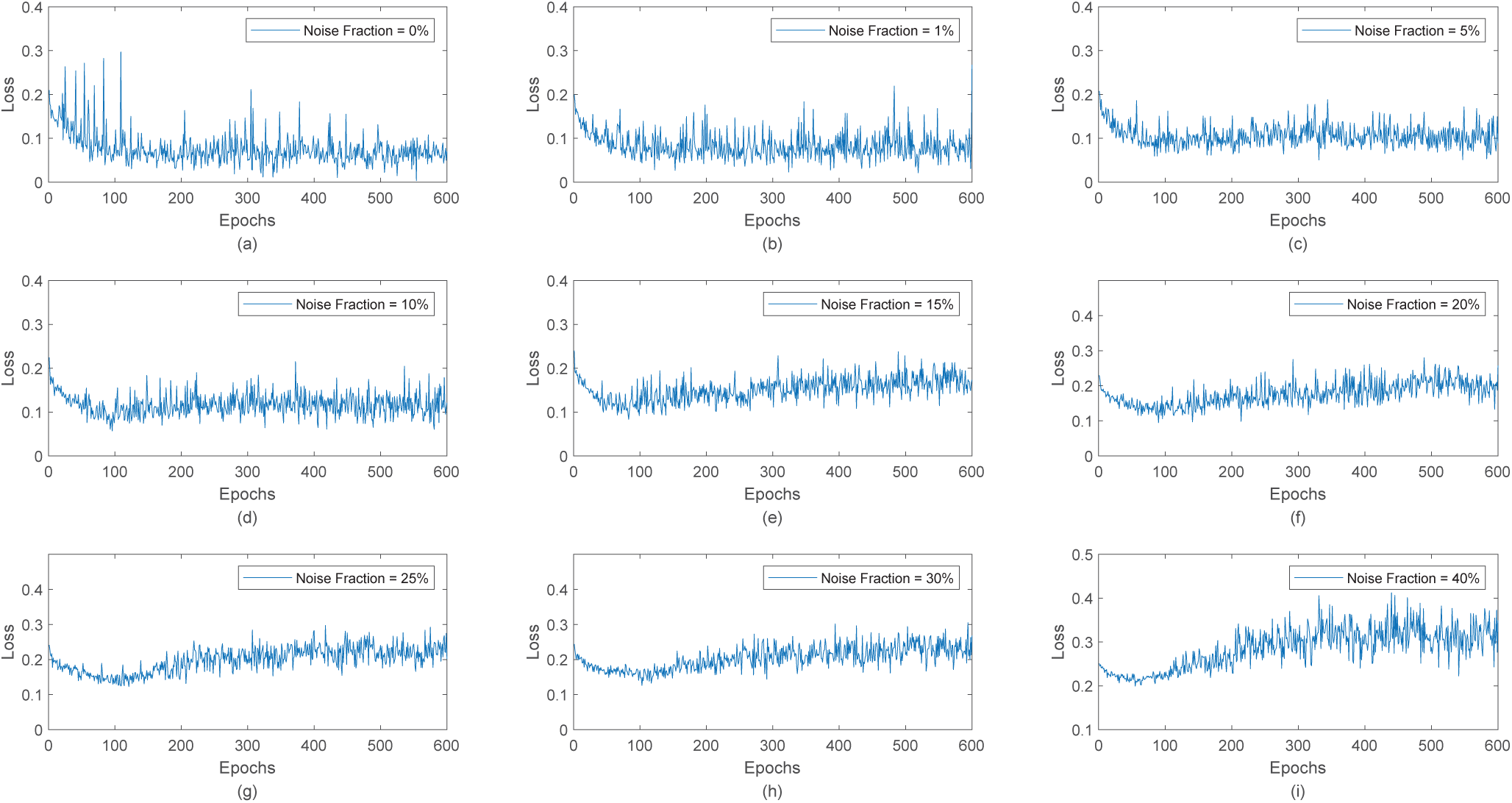
The loss functions of the proposed model under different noise fractions range from 0 to 40%.

In case of no other pneumonias, we can see from Table 4 that the number of the noisy labels in the training set has an effect on the model performance. When the noise fractions are 1%, 5% and 10%, the accuracy of the model in the testing set is down to 89.06%, 85.82% and 83.91%, respectively. Once the noise fraction is 15%, the accuracy is below 80%. These results show that the proposed AI model has an ability to classify COVID-19 from healthy subjects, and is robust to noisy label problem. In case of the healthy objects are hybrid with other pneumonias, we can see from Table 5 that the existence of the noisy labels in the training set will degrade the model performance. When the noise fractions are 1%, 5% and 10%, the accuracy of the model in the testing set is down to 84.89%, 82.61% and 79.11%, respectively. These results show that the proposed AI model has an ability to classify COVID-19 from other hybrid subjects, and is also robust to the case of a small part of the samples in the training set marked incorrectly.

As for the other three metrics including sensitivity, specificity and accuracy, we can see from Table 4 and Table 5 that the simulation results are similar. In order to better show the effect of noisy labels, we plot Fig. 7 with the averages of the accuracy, sensitivity, specificity and AUC values in the testing set with the propose method under different noise fractions. The *blue* line is the case of the classification of COVID-19 from healthy subjects when the *red* line is the the classification of COVID-19 from the other hybrid (healthy, bacterial pneumonia and SARS) subjects. We can see from this figure that as the noise fraction increases, all the four evaluation metrics decrease correspondingly. We can see from the figures in Fig. 7 that the blue line is higher than the red line in most cases, indicating that there exists some correlation property between the COIVD-19 and other pneumonia in chest CT image.

**Fig. 7.**
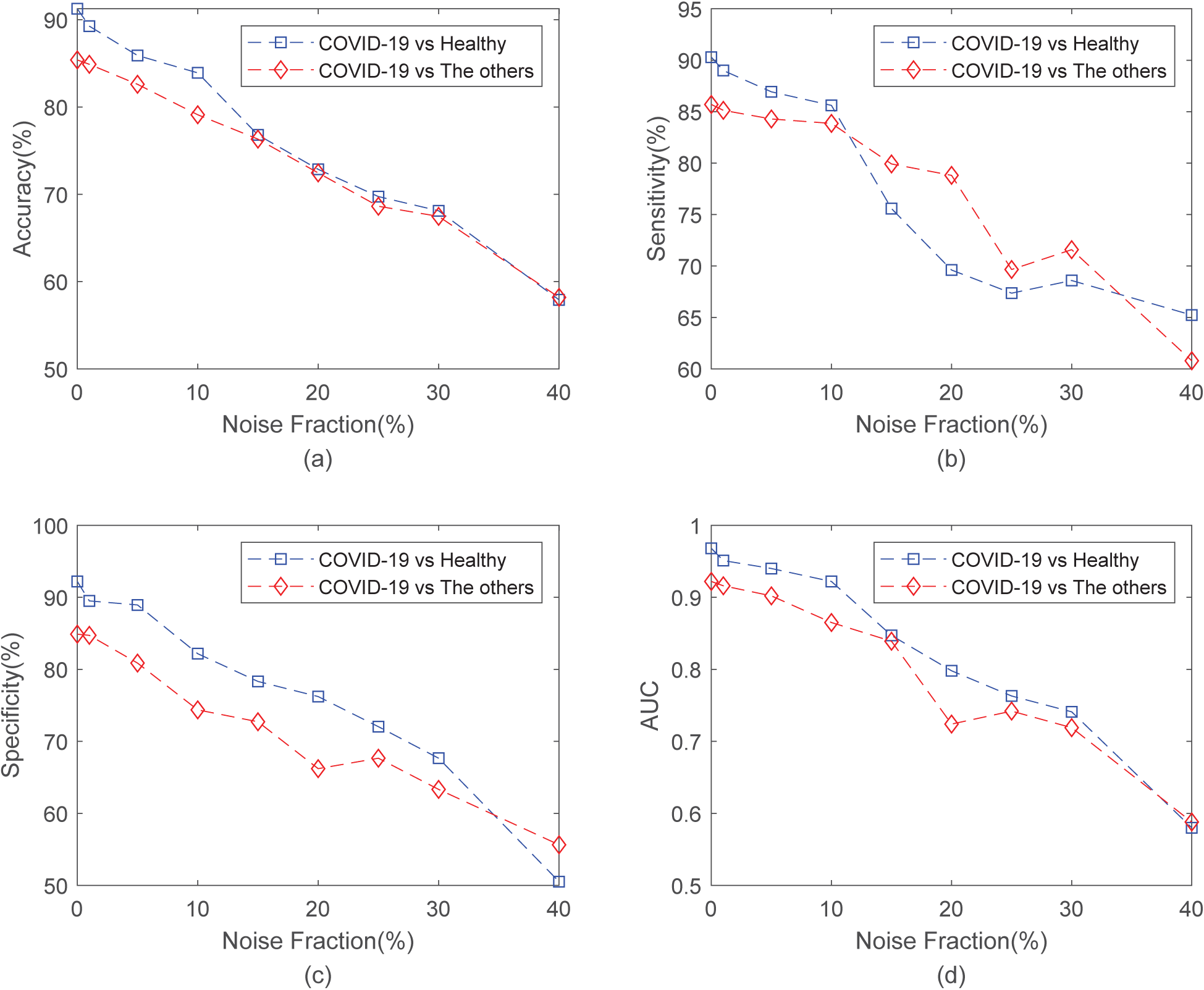
Performance of our model training with different percentage of the noise label. Accuracy, sensitivity, specificity and AUC are shown in (a), (b), (c) and (d), respectively.

## Discussions

### The operation of DA

We built an effective dataset of 1042 samples consisting of 521 COVID-19, 397 normal samples, 76 bacterial pneumonia and 48 SARS by searching available data on the Internet and selected those high-quality cases. Since the dataset is still small, we explore data augmentation technique to generate more training samples. Simulation results have proven the validity of these data augmentation operations in the proposed AI model.

### The power of AI

We present an AI method based on deep learning model by adopting ShuffleNet V2 network as the backbone for diagnosis of COVID-19 with the consideration of the sample amount, the detection accuracy and the training speed. Besides, we tested the effect of noisy labels in the training set on the model. Experimental results have shown that the proposed AI diagnosis model can classify a COVID-19 case from a heathy one or distinguish COVID-19 from other cases. The AI-based model is robust when part of the training set is labeled incorrectly. In the classification of COVID-19 from heathy subject, the average AUC of the proposed model is 0.9689 with [95%CI: 0.9308, 1] in case of no noisy labels. In the classification of COVID-19 from other hybrid cases, the average AUC of the proposed model is 0.9222 with [95%CI: 0.8418, 1] if there are no noisy labels. These results show that the AI methods based on deep learning model has an ability in assisting medical workers for more efficient diagnosis of COVID-19.

### Future Directions

There are two considerations in our future research. One is to continually add new high-quality samples into the dataset by keeping an eye on new public data sources or making a cooperation with those hospitals with valuable samples. The other consideration is to find a way to extract efficient features from chest CT images as input of the AI model for the improvement of model performance.

## Data Availability

The CT images are obtained from the internet, including the two proposed papers and a regular medical website.

https://arxiv.org/abs/2003.13865

https://arxiv.org/abs/2003.11597

https://www.sirm.org/category/senza-categoria/covid-19/

## Funding

This work was supported by NSFC (No. 61772234).

## Compliance

**Compliance of interest** None.

## Notes

### Competing Interest Statement

The authors have declared no competing interest.

